# Elevated α-synuclein aggregate levels in the urine of patients with isolated REM sleep behavior disorder and Parkinson’s disease

**DOI:** 10.1101/2025.01.14.25320280

**Authors:** Laura Müller, Pelin Özdüzenciler, Charlotte Schedlich-Teufer, Aline Seger, Hannah Jergas, Gereon R. Fink, Dieter Willbold, Michael Sommerauer, Michael T. Barbe, Gültekin Tamgüney

## Abstract

Parkinson’s disease is a neurodegenerative disorder characterized by α-synuclein aggregation in neurons. Recent advances suggest α-synuclein aggregates could serve as a biomarker for Parkinson’s disease and related synucleinopathies. This study used surface-based fluorescence intensity distribution analysis (sFIDA) to measure α-synuclein aggregates in urine. Patients with Parkinson’s disease and isolated REM sleep behavior disorder, a precursor to Parkinson’s disease, had elevated concentrations compared to healthy controls. Sensitivity and specificity were 83% and 65% for Parkinson’s disease versus controls and 89% and 62% for isolated REM sleep behavior disorder versus controls. The findings highlight sFIDA’s potential for diagnosing synucleinopathies.

## INTRODUCTION

Parkinson’s disease (PD) is a progressive neurodegenerative disease characterized by the pathological aggregation of α-synuclein in neurons.^1, 2^ The diagnosis of PD predominantly depends on clinical evaluation and brain imaging.^3^ Efforts to identify clinical and biological markers of disease heterogeneity and progression in PD are ongoing, yet a quantitative biomarker remains unavailable.^4^ An emerging biomarker for PD is aggregated α-synuclein, found in cerebrospinal fluid (CSF) and other biosamples.^5^ Importantly, patients with isolated rapid eye movement sleep behavior disorder (iRBD), a prodrome of PD and related synucleinopathies, also accumulate pathological α-synuclein aggregates in the nervous system.^6^

Quantifying α-synuclein aggregates in biosamples is challenging due to low concentrations and excess α-synuclein monomers.^7^ To address these challenges, we recently adapted surface-based fluorescence intensity distribution analysis (sFIDA) to measure α-synuclein aggregates in CSF and stool and demonstrated that sFIDA is a highly sensitive assay for quantifying α-synuclein aggregates in biosamples.^8, 9^ sFIDA utilizes the Syn211 antibody, which specifically targets amino acids 121–125 of α-synuclein to capture α-synuclein species. Since the Syn211 antibody recognizes only a single linear epitope on α-synuclein, its use as the detection antibody ensures precise quantification of aggregated α-synuclein species exclusively, as monomers with a single linear epitope are captured but remain undetected. Labeling the detection antibody with fluorophores and imaging the surface using confocal microscopy achieves near single particle sensitivity.

Considering that cellular components from the central and peripheral nervous systems can be released and distributed throughout the body and bloodstream, we hypothesized that aggregated α-synuclein species might likewise be detectable in the urine of patients with PD and iRBD.^4, 8-10^ The aim of this study was to detect and quantify α-synuclein aggregates in urine and, more importantly, to demonstrate their potential application in the diagnosis of patients with synucleinopathies.

## METHODS

### Participants

PD and iRBD patients and healthy controls (HC) were enrolled at the Department of Neurology of the University Hospital Cologne and diagnosed based on established criteria.^9^ Urine samples were collected between July 2020 and September 2021 and stored at −80 °C. All patient data and samples were pseudonymized. The study was approved by the research ethics committees of the participating institutions. Participants provided written informed consent.

### Sample preparation

Urine samples were thawed at 4 °C for 30 min, transferred to tubes coated with 3% bovine serum albumin (BSA, Applichem), and adjusted to 3% BSA and 1× Halt protease inhibitor cocktail (Thermo Fisher Scientific). After centrifugation at 4000 × g for 30 min at 4 °C, the supernatants were transferred to BSA-coated centrifugal concentrators (Sartorius) with a 10 kDa cutoff and concentrated 10-fold by centrifugation at 12,000 × g for 15−30 min at 4 °C. Concentrated samples were transferred to BSA-coated tubes and stored at −80 °C.

### sFIDA protocol

To capture α-synuclein aggregates, a 384-well glass-bottom microplate (Thermo Fisher Scientific) was incubated with 7.5 µg/mL of the Syn211 antibody (Santa Cruz Biotechnology) in 1× phosphate-buffered saline overnight at 4 °C. Subsequently, the plate was washed five times with 80 µL Tris-buffered saline (TBS) with 0.05% Tween 20 (TBS-T), and then five times with TBS. To block free binding sites, the wells were incubated with 0.5% fat-free dried milk powder in TBS with 0.03% ProClin (Sigma Aldrich, TBS-ProClin) for 3 h at room temperature. Next, the wells underwent five washes with 80 µL of TBS-T and five washes with TBS. The α-synuclein-coated silica nanoparticles (SiNaPs), which functioned as a standard for protein quantification, were prepared as previously described^8, 9, 11^ and diluted with 0.1% BSA and 0.05% Tween20 in TBS-ProClin. Four replicates of 20 µL of each SiNaP dilution or 20 µL of the 10× concentrated urine samples were added to the wells and incubated for 1 h at room temperature. Subsequently, the plate was washed five times with TBS. To detect α-synuclein aggregates, the wells were incubated with the fluorescent Syn211-CF633 and Syn211-CF488A antibodies,^9^ each at 0.625 µg/mL, in TBS-T containing 0.1% BSA. Finally, the plate was washed five times with TBS and the buffer exchanged to TBS-ProClin for measurement. An automated microtiter plate washer (405 LS Microplate Washer, BioTek) carried out all washing steps.

### Image-data acquisition and analysis

Images were captured using an IN Cell Analyzer 6500HS (GE Healthcare) with a 40× objective through two-channel confocal fluorescence imaging. A total of 25 images per well, each measuring 2040 × 2040 pixels, were captured for each channel. sFIDAta version 2.2.7 was utilized for the automated detection and elimination of images with artifacts, and for quantifying colocalized pixels in both channels (sFIDA readout).^9^ To minimize background, a cutoff was established as the gray value at which 0.01% of all pixels in images of the buffer control were positive. Only pixels with gray values above the cutoff were analyzed. The limit of detection (LOD) was defined as the sFIDA readout of the buffer control plus three standard deviations. A calibration curve was fitted using sFIDA readouts of dilutions of SiNaPs of known concentration and second order polynomial regression. This calibration curve was used to calculate a concentration for each sFIDA readout.

## RESULTS

### Descriptive analysis of the patient and control groups

We collected urine samples of 93 PD and 72 iRBD patients and 52 healthy controls (HC). Demographic, clinical, and statistical information for these cohorts is available in Tables 1 and S1 and Fig S1. There was a gender bias towards men in the patient cohorts, and a slight bias towards women in the control group. HCs were on average 10–12 years younger than patients. We did not observe significant differences in the duration of education. The cognitive performance (DemTect score) of HCs was significantly higher than that of patients.^12^ PD patients scored significantly higher than iRBD patients on the Movement Disorder Society’s Unified Parkinson’s Disease Rating Scale Part III.^13^ On average, PD patients scored 3.0 ± 0.9 on the Hoehn and Yahr scale and improved 42 ± 19% on the levodopa challenge test.^14^ PD patients scored significantly higher on the Non-Motor Symptoms Scale for PD than iRBD patients or HCs.^15^ Based on the Cleveland Clinic Constipation Scoring System, PD patients were significantly more constipated than iRBD patients and HCs.^16^ PD patients also suffered more from hyposmia, reflected in a significantly lower olfactory testing score, than iRBD patients or HCs. In addition, PD patients scored significantly higher in the screening questionnaire for parkinsonism compared to iRBD patients and HCs.^17^ The results of the RBD screening questionnaire also showed significant differences between iRBD and PD patients or HCs, as well as between PD patients and HCs.^18^

**Table 1.** Demographic and clinical information on patients and controls that donated urine samples.

|  | Group |  |  | *p* value |  |  |
| --- | --- | --- | --- | --- | --- | --- |
|  | PD | iRBD | HC | PD vs.<br>HC | PD vs.<br>iRBD | iRBD vs.<br>HC |
| Number | 93 | 72 | 52 | N/A | N/A | N/A |
| Female [number (percentage)] | 29 (31.2) | 10 (13.9) | 31 (59.6) | $p < 0.001$ | $p < 0.01$ | $p < 0.0001$ |
| Age [years ± SD] | 64.3 ± 9.7 | 66.3 ± 6.4 | 54.8 ± 16.8 | $p < 0.01$ | n.s. | $p < 0.001$ |
| Education [years ± SD] | 15.4 ± 4.1 | 15.9 ± 4.0 | 16.0 ± 3.3 | n.s. | n.s. | n.s. |
| Disease duration [years ± SD] | 9.0 ± 5.8 | 7.4 ± 5.4 | N/A | N/A | n.s. | N/A |
| DemTect [score ± SD] | 14.0 ± 3.1 | 14.8 ± 2.3 | 16.1 ± 2.0 | $p < 0.0001$ | n.s. | $p < 0.01$ |
| MDS-UPDRS III [score ± SD] | 23.7 ± 14.6 | 4.0 ± 2.7 | N/A | N/A | $p < 0.0001$ | N/A |
| Non-Motor Symptoms Scale<br>[score ± SD] | 30.3 ± 22.8 | 16.2 ± 18.0 | 11.0 ± 11.0 | $p < 0.0001$ | $p < 0.0001$ | n.s. |
| CCCSS [score ± SD] | 4.1 ± 4.0 | 2.8 ± 2.7 | 2.4 ± 2.5 | n.s. | n.s. | n.s. |
| Olfactory testing [score ± SD] | 5.4 ± 2.5 | 6.6 ± 2.7 | 10.3 ± 1.8 | $p < 0.0001$ | $p < 0.05$ | $p < 0.0001$ |
| Screening questionnaire for<br>parkinsonism [score ± SD] | 5.6 ± 2.2 | 0.3 ± 0.8 | 0.3 ± 0.7 | $p < 0.0001$ | $p < 0.0001$ | n.s. |
| RBD screening questionnaire<br>[score ± SD] | 4.9 ± 3.0 | 9.0 ± 2.7 | 2.1 ± 2.2 | $p < 0.0001$ | $p < 0.0001$ | $p < 0.0001$ |
| Hoehn and Yahr [score ± SD] | 3.0 ± 0.9 | N/A | N/A | N/A | N/A | N/A |
| Improvement of MDS-UPDRS<br>III on Levodopa challenge test<br>[% ± SD] | 42 ± 19 | N/A | N/A | N/A | N/A | N/A |

PD = Parkinson's disease; RBD = rapid eye movement sleep behavior disorder; HC = healthy control; SD = standard deviation; MDS-UPDRS III = Movement Disorder Society's Unified Parkinson's Disease Rating Scale Part III; CCCSS = Cleveland Clinic Constipation Scoring System; N/A = not available; n.s. = not significant.

### sFIDA measurements of α-synuclein-coated silica nanoparticles (SiNaPs)

To establish a standard for signal calibration, we utilized dilutions of α-synuclein-coated SiNaPs ranging from 0.1 to 3200 fM (Fig 1A).^11^ The mean intra-assay coefficient of variation (CV%) was determined based on the sFIDA readout of four replicates of each dilution of the SiNaPs standard, yielding a value of 19.9% (Table S2). The sFIDA readout of 3200 fM SiNaPs without a capture antibody was reduced by 99.7%. Based on the buffer control, the limit of detection (LOD) was determined to be 0.74 fM.

**Figure 1.**
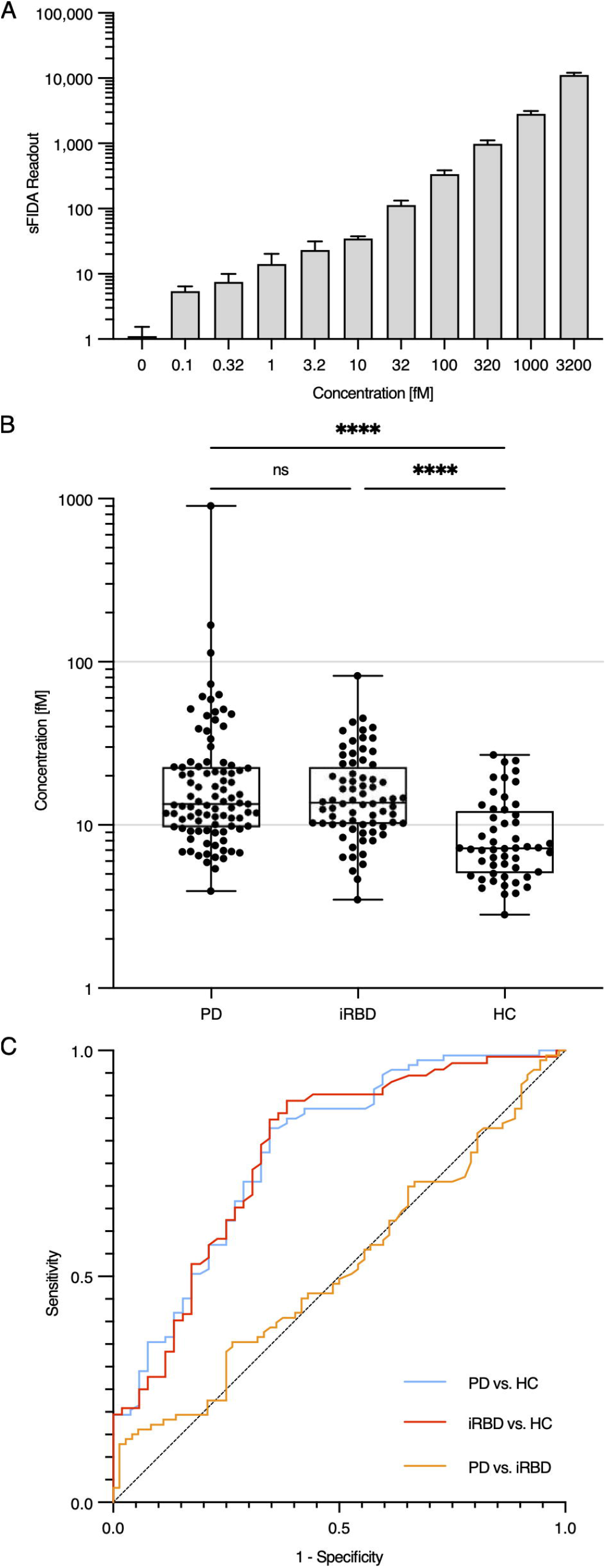
Concentration of α-synuclein aggregates in urine samples and receiver operating characteristics (ROC) curves. (A) sFIDA measurements of dilutions of α-synuclein-coated silica nanoparticles (SiNaPs) were utilized as a standard for signal calibration. (B) The median concentration of α-synuclein aggregates in 10× concentrated urine of patients with Parkinson’s disease (PD, 13.5 fM) and isolated rapid eye movement sleep behavior disorder (iRBD, 13.7 fM) was significantly (\*\*\*\**p* < 0.0001) higher than in the urine of the healthy controls (HC, 7.2 fM). sFIDA readouts were converted to concentrations using sFIDAta version 2.2.7. Significance was determined using GraphPad Prism Version 10.4.0 and the Kruskal-Wallis test (ns = non-significant). (C) A comparison of PD patients with healthy controls revealed a sensitivity of 83% at a specificity of 65% with an area under the curve (AUC) of 0.7763 (*p* < 0.0001). A comparison between iRBD patients and healthy controls yielded a sensitivity of 89% and a specificity of 62% with an AUC of 0.7743 (*p* < 0.0001). The ROC analyses were performed using GraphPad Prism Version 10.4.0. The optimal combination of sensitivity and specificity was calculated with a maximized Youden’s index.

### α-Synuclein aggregate concentrations are elevated in the urine of PD and iRBD patients

We employed sFIDA to measure α-synuclein aggregate concentrations in the urine of HCs and patients with PD and iRBD. We used SiNaPs to calibrate the sFIDA readouts and calculate the concentrations of α-synuclein aggregates (Fig 1B, Table S1). The median concentration of α-synuclein aggregates in 10× concentrated urine samples from PD (13.5 fM) and iRBD patients (13.7 fM) was significantly elevated (*p* < 0.0001) compared to those from HCs (7.2 fM) using the Kruskal-Wallis test. We observed no significant difference between PD and iRBD patients. The receiver operating characteristic (ROC) curves (Fig 1C) revealed an 83% sensitivity and 65% specificity for PD patients versus HCs, and an 89% sensitivity and 62% specificity for iRBD patients versus HCs. We observed no correlation between α-synuclein aggregate concentrations in urine and other disease-relevant scores (Table S3).

## DISCUSSION

We demonstrated that α-synuclein aggregate concentrations are significantly elevated in the urine of PD and iRBD patients compared to HCs. Notably, the concentrations in iRBD patients matched those we observed in PD patients. These findings corroborate and expand on a previous study reporting higher (*p* < 0.05) urinary α-synuclein aggregate concentrations in 21 PD patients compared to 11 HCs.^19^ While sFIDA for urine is nearly as sensitive as the seed amplification assay for detecting α-synuclein aggregates in CSF samples of various synucleinopathies, it is less specific.^4^ We also observed α-synuclein aggregates in the urine of some control subjects, possibly because sFIDA cannot differentiate between seeding-competent fibrillar aggregates, which are neurotoxic, and non-fibrillar, seeding-incompetent forms that may be present in healthy individuals.^20^ Alternatively, some control subjects may be in the clinical prodromal stage of PD. The origin of urinary α-synuclein aggregates remains uncertain but may be linked to blood or peripheral nerves.^5^ In summary, our results demonstrate that pathological α-synuclein aggregates are excreted in urine and can be quantified using sFIDA, highlighting its potential for diagnosing synucleinopathies in their early stages.

## Data Availability

All data produced in the present work are contained in the manuscript.

## Acknowledgements

Parts of this study were supported by the Else Kröner-Fresenius-Stiftung (2019_EKES.02, MS), Koeln Fortune (HJ), FMMED (HJ), and EIT Health (HJ), the Deutsche Forschungsgemeinschaft (DFG, German Research Foundation – Project-ID 431549029 – SFB 1451, GRF), and the Ilselore-Luckow-Stiftung (GT).

## Author Contributions

MS, MTB, GRF, DW, and GT supervised the project. CST, HJ, MTB, and MS recruited and clinically assessed patients and control subjects, and collected samples. LM and GT developed the sFIDA assay for urine samples. LM and PÖ performed all experiments. LM and GT analyzed the data. LM, MS, MTB, and GT wrote the manuscript. All authors critically revised the manuscript.

## Data Availability

The authors confirm that the data supporting the findings of this study are available within the article and its supplementary materials.

