## Supplementary Materials for "Elevated α-synuclein aggregate levels in the urine of patients with isolated REM sleep behavior disorder and Parkinson’s disease"

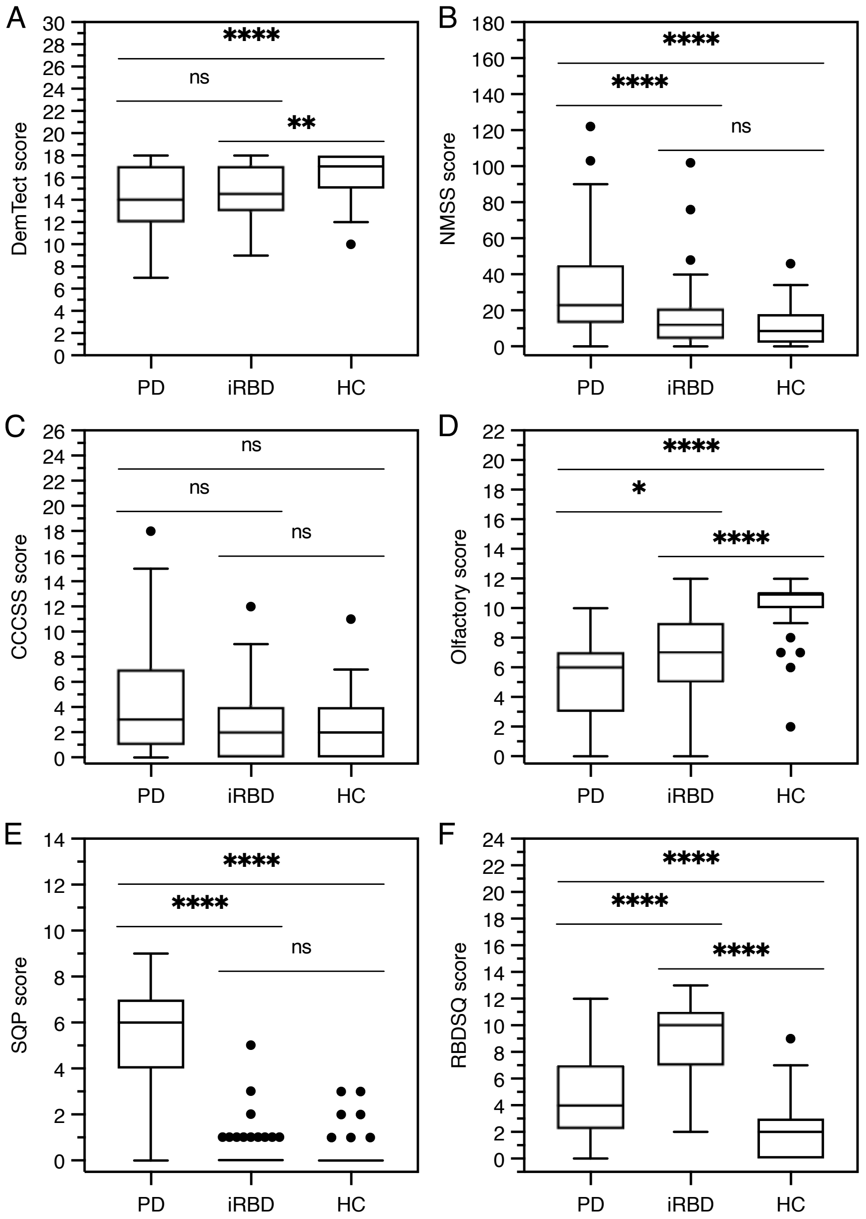


**Figure S1:** Characteristics for PD and iRBD patients and healthy controls based on their performance in various tests and screening questionnaires. (A) The DemTect score showed that the cognitive performance of healthy controls (16.1 ± 2.0) was significantly higher ($p < 0.0001$) than that of PD (14.0 ± 3.3) and iRBD patients (14.8 ± 2.3, $p < 0.01$). (B) Based on the Non-Motor Symptoms Scale (NMSS), the performance of PD patients (30.3 ± 22.8) was significantly worse ($p < 0.0001$) than that of iRBD patients (16.2 ± 18.0) and healthy controls (11.0 ± 11.0). (C) Based on the Cleveland Clinic Constipation Scoring System (CCCSS), there was no significant difference in constipation of PD patients (4.1 ± 4.0), iRBD patients (2.8 ± 2.7), and healthy controls (2.5 ± 2.5). (D) The olfactory performance of PD patients (5.4 ± 2.5) was significantly worse ($p < 0.05$) than that of iRBD patients (6.6 ± 2.7). That of PD and iRBD patients was significantly worse ($p < 0.0001$) than that of healthy controls (10.3 ± 1.8), based on correctly identified Sniffin' sticks. (E) PD patients (4.9 ± 3.0) scored significantly higher ($p < 0.0001$) than iRBD patients (0.3 ± 0.8) and healthy controls (0.3 ± 0.7) on the Screening Questionnaire for Parkinsonism (SQP). (F) The RBD Screening Questionnaire (RBDSQ) revealed significant differences ($p < 0.0001$) between iRBD (9.0 ± 2.7) and PD patients (4.9 ± 3.0) or healthy controls (2.1 ± 2.2), and between PD patients and healthy controls. Significance was determined using GraphPad Prism Version 10.4.0 and the Kruskal-Wallis test (ns = non-significant, *$p < 0.05$, **$p < 0.01$, ****$p < 0.0001$).

**Table S1.** Demographic and clinical information and concentrations of α-synuclein aggregates in urine

| Sample | Diagnosis | Sex | Age range at sampling [yrs] | Age range at onset [yrs] | Disease duration [yrs] | Education [yrs] | CCCSS score | DemTect score | NMSS score | MDS-UDPRS III score | Olfactory testing score | Hoehn and Yahr score | Levodopa challenge test % change | DaTSCAN score | Screening questionnaire PD score | RBDSQ score | α-Synuclein aggregate concentration [fM] |
|---|---|---|---|---|---|---|---|---|---|---|---|---|---|---|---|---|---|
| 1 | PD | Male | 71-75 | 56-60 | 13 | 18 | 9 | 13 | 37 | 22 | 4 | 4 | 69 | + | 5 | 7 | 7.32 |
| 2 | PD | Male | 66-70 | 51-55 | 14 | 10 | 10 | 13 | 17 | 27 | 6 | 3 | 33 | N/A | 6 | 10 | 3.02 |
| 3 | PD | Male | 56-60 | 41-45 | 11 | 12 | 3 | 17 | 51 | 35 | 7 | 4 | 53 | + | 5 | 8 | 4.41 |
| 4 | PD | Male | 71-75 | 61-65 | 10 | 17 | 1 | 11 | 67 | 65 | 8 | 5 | 24 | + | 9 | 6 | 2.44 |
| 5 | PD | Male | 46-50 | 41-45 | 5 | 12 | 0 | 14 | 103 | 21 | 3 | 3 | 44 | N/A | 6 | 2 | 4.79 |
| 6 | PD | Male | 71-75 | 56-60 | 15 | 19 | 6 | 14 | 54 | 28 | 5 | 5 | 47 | N/A | 4 | 5 | 4.03 |
| 7 | PD | Male | 51-55 | 46-50 | 4 | 16 | 0 | 18 | 20 | 19 | 2 | 2 | 47 | + | 7 | 0 | 5.12 |
| 8 | PD | Female | 71-75 | 46-50 | 22 | 12 | 4 | 9 | 67 | 20 | 9 | 3 | 48 | N/A | 9 | 4 | 1.82 |
| 9 | PD | Male | 56-60 | 51-55 | 4 | 16 | 2 | 15 | 9 | 33 | 7 | 5 | 32 | N/A | 7 | 5 | 4.67 |
| 10 | PD | Male | 61-65 | 46-50 | 12 | 20 | 0 | 18 | 9 | 10 | 2 | 2 | 46 | + | 8 | 3 | 6.30 |
| 11 | PD | Male | 76-80 | 76-80 | 2 | 8 | 3 | 14 | 4 | 17 | 3 | 2 | - | N/A | 1 | 0 | 6.14 |
| 12 | PD | Female | 76-80 | 61-65 | 15 | 11 | 18 | 17 | 48 | 76 | 2 | 5 | 43 | N/A | 6 | 4 | 5.90 |
| 13 | PD | Male | 66-70 | 56-60 | 10 | 18 | 9 | 16 | 4 | 17 | 3 | 4 | 63 | N/A | 5 | 2 | 4.94 |
| 14 | PD | Female | 76-80 | 66-70 | 14 | 13 | 11 | 13 | 34 | 25 | 7 | 4 | 53 | + | 7 | 1 | 2.41 |
| 15 | PD | Female | 51-55 | 31-35 | 17 | 19 | 1 | 12 | 27 | 23 | 4 | 4 | 58 | N/A | 5 | 7 | 3.76 |
| 16 | PD | Female | 71-75 | 56-60 | 14 | 17 | 9 | 10 | 21 | 50 | 2 | 5 | 32 | N/A | 9 | 6 | 2.22 |
| 17 | PD | Male | 61-65 | 51-55 | 9 | 13 | 1 | 12 | 12 | 15 | 4 | 2 | 64 | + | 7 | 4 | 5.14 |
| 18 | PD | Male | 71-75 | 61-65 | 12 | 12 | 3 | 11 | 21 | 8 | 0 | 2 | 32 | N/A | 2 | 3 | 1.32 |
| 19 | PD | Male | 81-85 | 71-75 | 9 | 12 | 0 | 9 | 46 | 23 | 8 | 4 | 38 | N/A | 7 | 5 | 1.08 |
| 20 | PD | Female | 46-50 | 36-40 | 11 | 14 | 2 | 11 | 16 | 22 | 7 | 2 | 46 | + | 5 | 2 | 1.13 |
| 21 | PD | Female | 56-60 | 51-55 | 5 | 12 | 0 | 15 | 4 | 30 | 9 | 3 | 32 | + | 7 | 5 | 3.89 |
| 22 | PD | Female | 56-60 | 51-55 | 9 | 17 | 3 | 17 | 16 | 16 | 7 | 2 | 42 | + | 7 | 2 | 0.66 |

| | | | | | | | | | | | | | | | | | |
|---|---|---|---|---|---|---|---|---|---|---|---|---|---|---|---|---|---|
| 23 | PD | Male | 56-60 | 36-40 | 20 | 12 | 7 | 18 | 15 | 30 | 3 | 3 | 47 | N/A | 5 | 3 | 2.26 |
| 24 | PD | Male | 56-60 | 36-40 | 18 | 15 | 7 | 12 | 42 | 41 | 6 | 3 | 40 | + | 5 | 8 | 2.12 |
| 25 | PD | Male | 56-60 | 56-60 | 3 | 17 | 13 | 18 | 46 | 7 | 4 | 2 | 48 | N/A | 7 | 9 | 1.19 |
| 26 | PD | Female | 56-60 | 56-60 | 0 | 20 | 2 | 18 | 52 | 16 | 1 | 2 | - | + | 0 | 11 | 0.95 |
| 27 | PD | Male | 66-70 | 61-65 | 6 | 20 | 1 | 17 | 10 | 5 | 6 | 2 | - | + | 4 | 2 | 1.10 |
| 28 | PD | Male | 76-80 | 71-75 | 2 | 13 | 1 | 11 | 21 | 35 | 6 | 3 | - | N/A | 6 | 8 | 0.98 |
| 29 | PD | Male | 56-60 | 51-55 | 9 | 21 | 4 | 14 | 56 | 22 | 1 | 3 | 49 | N/A | 8 | 5 | 1.35 |
| 30 | PD | Male | 56-60 | 51-55 | 5 | 19 | 4 | 18 | 28 | 33 | 6 | 3 | - | + | 7 | 12 | 1.36 |
| 31 | PD | Male | 61-65 | 61-65 | 2 | 15 | 6 | 12 | 28 | 42 | 5 | 4 | 43 | N/A | 9 | 8 | 0.70 |
| 32 | PD | Male | 41-45 | 36-40 | 1 | 22 | 2 | 18 | 7 | 11 | 10 | 2 | - | + | 2 | 3 | 2.43 |
| 33 | PD | Male | 66-70 | 41-45 | 21 | 15 | 5 | 18 | 14 | 11 | 6 | 2 | 0 | + | 4 | 4 | 1.50 |
| 34 | PD | Female | 56-60 | 46-50 | 10 | 13 | 12 | 13 | 27 | 19 | 6 | 3 | 43 | + | 8 | 7 | 1.54 |
| 35 | PD | Male | 71-75 | 66-70 | 5 | 12 | 2 | 8 | 30 | 42 | 9 | 4 | 25 | + | 4 | 1 | 0.82 |
| 36 | PD | Male | 56-60 | 51-55 | 4 | 15 | 2 | 11 | 42 | 10 | 8 | 3 | 69 | N/A | 7 | 6 | 1.30 |
| 37 | PD | Female | 66-70 | 56-60 | 13 | 8 | 11 | 11 | 25 | 22 | 8 | 3 | 44 | N/A | 6 | 4 | 1.40 |
| 38 | PD | Male | 51-55 | 41-45 | 14 | 9 | 12 | 7 | 30 | 50 | 6 | 4 | 45 | N/A | 8 | 6 | 0.92 |
| 39 | PD | Male | 66-70 | 61-65 | 4 | 13 | 6 | 9 | 65 | 21 | 3 | 3 | - | + | 9 | 6 | 2.11 |
| 40 | PD | Male | 76-80 | 71-75 | 7 | 18 | 7 | 10 | 10 | 19 | 7 | 3 | - | + | 8 | 12 | 0.62 |
| 41 | PD | Male | 51-55 | 46-50 | 5 | 16 | 1 | 14 | 1 | 5 | 8 | 2 | 54 | N/A | 3 | 5 | 1.64 |
| 42 | PD | Female | 46-50 | 41-45 | 5 | 15 | 5 | 18 | 13 | 9 | 8 | 3 | 0 | N/A | 7 | 3 | 1.15 |
| 43 | PD | Female | 61-65 | 46-50 | 15 | 25 | 4 | 17 | 22 | - | 5 | 4 | 22 | N/A | 5 | 11 | 0.68 |
| 44 | PD | Male | 41-45 | 36-40 | 4 | 20 | 3 | 14 | 17 | 18 | 6 | 3 | - | + | 6 | 1 | 11.36 |
| 45 | PD | Male | 66-70 | 56-60 | 11 | 16 | 3 | 10 | 20 | 21 | 6 | 3 | 52 | N/A | 5 | 5 | 0.95 |
| 46 | PD | Male | 61-65 | 51-55 | 10 | 13 | 8 | 14 | 21 | 13 | 10 | 2 | 31 | + | 7 | 8 | 0.39 |
| 47 | PD | Male | 71-75 | 61-65 | 6 | 12 | 3 | 10 | 26 | 48 | 4 | 3 | 16 | + | 7 | 1 | 2.16 |
| 48 | PD | Female | 76-80 | 61-65 | 14 | 20 | 6 | 18 | 23 | 30 | 4 | 3 | 29 | N/A | 7 | 2 | 0.80 |
| 49 | PD | Male | 76-80 | 56-60 | 16 | 12 | 11 | 13 | 90 | 31 | 6 | 4 | 30 | N/A | 6 | 9 | 1.15 |
| 50 | PD | Male | 51-55 | 51-55 | 0 | 30 | 0 | 17 | 2 | 5 | 7 | 2 | - | + | 1 | 3 | 1.26 |
| 51 | PD | Male | 76-80 | 61-65 | 12 | 18 | 3 | 13 | 26 | 26 | 7 | 3 | -2 | N/A | 4 | 8 | 0.54 |
| 52 | PD | Male | 61-65 | 51-55 | 12 | 22 | 9 | 16 | 20 | 25 | 7 | 2 | 93 | N/A | 8 | 4 | 1.86 |

| | | | | | | | | | | | | | | | | | |
|---|---|---|---|---|---|---|---|---|---|---|---|---|---|---|---|---|---|
| 53 | PD | Male | 81-85 | 71-75 | 10 | 22 | 8 | 18 | 36 | 54 | 5 | 4 | - | N/A | 6 | 7 | 1.29 |
| 54 | PD | Male | 61-65 | 51-55 | 9 | 18 | 3 | 15 | 44 | 6 | 2 | 2 | 33 | + | 5 | 6 | 1.33 |
| 55 | PD | Female | 66-70 | 61-65 | 7 | 14 | 0 | 12 | 18 | 13 | 6 | 3 | 37 | N/A | 8 | 4 | 1.73 |
| 56 | PD | Female | 71-75 | 61-65 | 13 | 8 | 3 | 12 | 19 | 63 | 4 | 5 | - | N/A | 7 | 7 | 1.28 |
| 57 | PD | Female | 76-80 | 46-50 | 27 | 13 | 1 | 18 | 67 | 30 | 2 | 4 | 57 | N/A | 9 | 7 | 1.10 |
| 58 | PD | Female | 61-65 | 46-50 | 14 | 23 | 7 | 18 | 11 | 40 | 7 | 5 | 9 | + | 6 | 4 | 16.76 |
| 59 | PD | Female | 71-75 | 46-50 | 26 | 16 | 2 | 9 | 36 | 25 | 2 | 3 | - | N/A | 5 | 4 | 0.98 |
| 60 | PD | Male | 56-60 | 56-60 | 2 | 14 | 1 | 14 | 29 | 41 | 9 | 3 | - | N/A | 4 | 3 | 2.31 |
| 61 | PD | Male | 66-70 | 56-60 | 10 | 14 | 0 | 18 | 24 | 24 | 0 | 2 | 61 | N/A | 7 | 4 | 1.46 |
| 62 | PD | Male | 71-75 | 66-70 | 5 | 18 | 0 | 18 | 14 | 22 | 7 | 3 | - | + | 3 | 4 | 1.18 |
| 63 | PD | Male | 56-60 | 46-50 | 8 | 15 | 1 | 14 | 13 | 10 | 9 | 2 | 55 | N/A | 3 | 2 | 3.38 |
| 64 | PD | Female | 51-55 | 41-45 | 9 | 16 | 2 | 18 | 15 | 9 | 10 | 3 | 69 | + | 7 | 3 | 0.65 |
| 65 | PD | Female | 56-60 | 41-45 | 12 | 12 | 2 | 18 | 122 | 26 | 7 | 2 | 72 | N/A | 8 | 7 | 0.98 |
| 66 | PD | Female | 66-70 | 51-55 | 17 | 12 | 0 | 17 | 19 | 19 | 8 | 4 | 32 | N/A | 2 | 8 | 1.18 |
| 67 | PD | Female | 61-65 | 56-60 | 7 | 20 | 8 | 13 | 59 | 21 | 8 | 3 | 63 | N/A | 8 | 11 | 0.69 |
| 68 | PD | Male | 56-60 | 46-50 | 8 | 13 | 10 | 12 | 39 | 22 | 4 | 3 | 31 | + | 6 | 6 | 0.77 |
| 69 | PD | Female | 71-75 | 71-75 | 1 | 17 | 2 | 15 | 15 | 16 | 8 | 3 | - | + | 6 | 9 | 1.12 |
| 70 | PD | Male | 46-50 | 36-40 | 10 | 24 | 1 | 15 | 22 | 16 | 9 | 2 | 67 | + | 6 | 3 | 1.06 |
| 71 | PD | Female | 66-70 | 56-60 | 13 | 11 | 7 | 15 | 51 | 28 | 4 | 3 | 42 | N/A | 7 | 6 | 0.59 |
| 72 | PD | Male | 51-55 | 51-55 | 2 | 18 | 12 | 14 | 50 | 17 | 3 | 2 | - | N/A | 8 | 6 | 2.26 |
| 73 | PD | Male | 76-80 | 56-60 | 18 | 17 | 2 | 12 | 36 | 59 | 3 | 5 | - | N/A | 7 | 5 | 2.02 |
| 74 | PD | Male | 51-55 | 41-45 | 7 | 16 | 7 | 9 | 51 | 21 | 8 | 3 | 35 | + | 6 | 2 | 0.75 |
| 75 | PD | Female | 56-60 | 51-55 | 2 | 15 | 2 | 18 | 12 | 6 | 6 | 3 | 73 | + | 1 | 1 | 0.69 |
| 76 | PD | Male | 51-55 | 41-45 | 10 | - | - | 7 | 79 | 22 | 7 | 3 | 48 | + | - | - | 90.28 |
| 77 | PD | Male | 56-60 | 56-60 | 3 | 10 | 0 | 12 | 55 | 36 | 2 | 3 | - | + | 1 | 1 | 1.01 |
| 78 | PD | Male | 51-55 | 46-50 | 6 | 18 | 6 | 13 | 0 | 5 | 6 | 2 | 0 | N/A | 3 | 1 | 1.18 |
| 79 | PD | Male | 51-55 | 36-40 | 11 | 14 | 7 | 8 | 10 | 31 | 4 | 4 | 49 | + | 5 | 5 | 0.69 |
| 80 | PD | Male | 71-75 | 71-75 | 1 | 14 | 15 | 15 | 62 | 23 | 1 | 3 | - | N/A | 5 | 1 | 0.64 |
| 81 | PD | Male | 66-70 | 51-55 | 11 | 12 | 0 | 14 | 51 | 13 | 7 | 2 | - | + | 4 | 2 | 0.92 |
| 82 | PD | Female | 66-70 | 51-55 | 14 | 12 | 3 | 15 | 57 | 31 | 8 | 4 | 34 | N/A | 9 | 10 | 0.90 |

| | | | | | | | | | | | | | | | | | |
|---|---|---|---|---|---|---|---|---|---|---|---|---|---|---|---|---|---|
| 83 | PD | Male | 71-75 | 66-70 | 6 | 13 | 1 | 15 | 13 | 12 | 5 | 3 | - | N/A | 7 | 9 | 1.20 |
| 84 | PD | Male | 56-60 | 46-50 | 10 | 13 | 3 | 14 | 29 | 13 | 3 | 2 | 47 | N/A | 7 | 2 | 0.92 |
| 85 | PD | Male | 71-75 | 66-70 | 6 | 8 | 2 | 9 | 6 | 42 | 6 | - | 54 | N/A | 4 | 1 | 1.83 |
| 86 | PD | Female | 61-65 | 56-60 | 6 | 16 | 0 | 13 | 13 | 10 | 2 | 2 | - | N/A | 2 | 6. | 2.06 |
| 87 | PD | Male | 71-75 | 61-65 | 7 | 19 | 0 | 18 | 18 | 29 | 3 | 3 | - | N/A | 5 | 1 | 2.07 |
| 88 | PD | Male | 51-55 | 51-55 | 0 | 13 | 1 | 17 | 12 | 11 | 3 | 2 | - | + | 4 | 11 | 1.40 |
| 89 | PD | Female | 56-60 | 56-60 | 1 | 14 | 3 | 13 | 24 | 6 | 8 | 2 | - | + | 5 | 4 | 1.71 |
| 90 | PD | Male | 76-80 | 71-75 | 6 | 13 | 0 | 17 | 11 | 14 | 6 | 3 | 44 | + | 2 | 1 | 0.91 |
| 91 | PD | Male | 61-65 | 56-60 | 3 | 22 | 1 | 14 | 13 | 7 | 7 | 2 | - | + | 2 | 3 | 2.08 |
| 92 | PD | Male | 61-65 | 61-65 | 1 | 18 | 0 | 13 | 12 | 4 | 2 | 2 | - | + | 3 | 3 | 1.50 |
| 93 | PD | Male | 76-80 | 76-80 | 3 | 10 | 1 | 13 | 31 | 17 | 1 | 2 | 14 | + | 4 | 3 | 2.28 |
| 94 | iRBD | Male | 56-60 | - | - | 21 | 2 | 17 | 6 | 1 | 11 | - | - | N/A | 0 | 6 | 1.02 |
| 95 | iRBD | Male | 71-75 | 56-60 | 12 | 3 | - | 16 | 1 | 4 | 11 | - | - | N/A | 2 | 6 | 1.33 |
| 96 | iRBD | Male | 61-65 | - | - | 16 | - | 14 | 4 | 5 | 11 | - | - | N/A | 0 | 6 | 1.47 |
| 97 | iRBD | Female | 56-60 | 51-55 | 4 | 20 | 4 | 17 | 14 | 0 | 6 | - | - | N/A | 0 | 10 | 1.42 |
| 98 | iRBD | Male | 76-80 | - | - | 14 | - | 17 | 15 | 4 | 8 | - | - | N/A | 0 | 10 | 1.13 |
| 99 | iRBD | Female | 66-70 | 46-50 | 17 | 12 | 3 | 13 | - | 8 | 5 | - | - | N/A | 0 | 9 | 2.40 |
| 100 | iRBD | Male | 61-65 | 61-65 | 0 | 16 | 2 | 17 | 3 | 5 | 10 | - | - | N/A | 0 | 8 | 0.64 |
| 101 | iRBD | Male | 56-60 | 51-55 | 4 | 16 | 0 | 13 | - | 6 | 9 | - | - | N/A | 0 | 10 | 0.98 |
| 102 | iRBD | Female | 61-65 | 61-65 | 3 | 21 | 0 | 15 | - | 3 | 3 | - | - | N/A | 0 | 7 | 1.82 |
| 103 | iRBD | Male | 61-65 | 61-65 | 1 | 15 | 4 | 11 | - | 2 | 7 | - | - | + | 0 | 8 | 1.35 |
| 104 | iRBD | Male | 66-70 | 56-60 | 10 | - | 0 | 14 | 12 | 5 | 10 | - | - | N/A | 0 | 12 | 8.19 |
| 105 | iRBD | Male | 66-70 | 51-55 | 14 | 17 | 0 | 12 | 4 | 5 | 7 | - | - | + | 0 | 11 | 1.25 |
| 106 | iRBD | Male | 71-75 | 71-75 | 2 | - | - | 12 | 2 | 4 | 6 | - | - | N/A | - | - | 0.47 |
| 107 | iRBD | Female | 61-65 | 56-60 | 5 | 16 | 1 | 17 | 6 | 3 | 5 | - | - | + | 0 | 9 | 0.80 |
| 108 | iRBD | Male | 71-75 | 61-65 | 10 | 11 | 2 | 14 | - | 5 | 9 | - | - | + | 0 | - | 1.30 |
| 109 | iRBD | Male | 71-75 | 71-75 | 3 | 18 | 2 | 14 | 8 | 7 | 3 | - | - | N/A | 0 | 7 | 0.35 |
| 110 | iRBD | Male | 66-70 | 61-65 | 5 | 14 | 0 | 17 | 9 | 4 | 8 | - | - | + | 0 | 12 | 0.87 |
| 111 | iRBD | Female | 66-70 | 61-65 | 4 | 15 | 5 | 18 | 2 | 3 | 9 | - | - | N/A | 0 | 5 | 3.82 |
| 112 | iRBD | Male | 56-60 | 56-60 | 3 | 19 | 3 | 9 | 22 | 5 | 7 | - | - | + | 0 | 9 | 1.04 |

| | | | | | | | | | | | | | | | | | |
|---|---|---|---|---|---|---|---|---|---|---|---|---|---|---|---|---|---|
| 113 | iRBD | Male | 71-75 | 66-70 | 9 | 18 | 0 | 17 | 1 | 5 | 7 | - | - | N/A | 0 | 11 | 1.42 |
| 114 | iRBD | Male | 51-55 | 36-40 | 15 | 21 | 0 | 11 | 0 | 8 | 12 | - | - | N/A | 0 | 5 | 1.27 |
| 115 | iRBD | Male | 66-70 | 66-70 | 3 | 15 | 8 | 12 | - | 13 | 7 | - | - | + | 0 | 11 | 1.85 |
| 116 | iRBD | Male | 51-55 | 46-50 | 5 | 12 | 5 | 12 | 48 | 3 | 8 | - | - | + | 5 | 13 | 0.90 |
| 117 | iRBD | Male | 71-75 | 71-75 | 2 | 16 | 0 | 15 | - | 6 | 7 | - | - | N/A | 0 | 10 | 1.03 |
| 118 | iRBD | Male | 66-70 | 61-65 | 5 | 15 | 3 | 18 | 21 | 7 | 6 | - | - | N/A | 0 | 10 | 1.01 |
| 119 | iRBD | Male | 66-70 | 61-65 | 7 | 20 | 2 | 13 | 31 | 8 | 1 | - | - | N/A | 1 | 5 | 2.93 |
| 120 | iRBD | Male | 66-70 | 46-50 | 17 | 13 | 0 | 17 | 2 | 10 | 7 | - | - | N/A | 0 | 10 | 1.35 |
| 121 | iRBD | Male | 66-70 | 56-60 | 11 | 17 | 0 | 17 | 3 | 4 | 9 | - | - | + | 0 | 10 | 4.52 |
| 122 | iRBD | Male | 61-65 | 46-50 | 15 | 31 | 4 | 14 | - | 5 | 9 | - | - | + | 1 | 9 | 2.37 |
| 123 | iRBD | Male | 66-70 | - | - | 15 | 0 | 18 | 1 | 4 | 7 | - | - | N/A | 0 | 5 | 1.03 |
| 124 | iRBD | Female | 61-65 | 61-65 | 1 | 19 | 12 | 14 | - | 0 | 4 | - | - | N/A | 0 | 8 | 0.63 |
| 125 | iRBD | Male | 71-75 | 51-55 | 20 | 15 | 6 | 13 | - | 4 | 8 | - | - | + | 1 | 8 | 1.42 |
| 126 | iRBD | Male | 66-70 | 61-65 | 6 | 16 | 5 | 13 | 23 | 1 | 3 | - | - | N/A | 0 | 11 | 2.82 |
| 127 | iRBD | Male | 66-70 | 66-70 | 3 | 22 | 3 | 14 | - | 3 | 3 | - | - | + | 1 | 10 | 2.06 |
| 128 | iRBD | Male | 71-75 | 71-75 | 2 | 12 | 2 | 11 | - | 5 | 5 | - | - | N/A | 0 | 7 | 1.90 |
| 129 | iRBD | Male | 56-60 | 51-55 | 5 | 13 | 9 | 14 | - | 7 | 5 | - | - | + | 3 | 11 | 1.03 |
| 130 | iRBD | Female | 76-80 | 66-70 | 11 | 16 | 1 | 15 | 4 | 5 | 10 | - | - | N/A | 0 | 12 | 1.01 |
| 131 | iRBD | Female | 56-60 | 51-55 | 4 | 16 | 1 | 15 | - | 4 | 10 | - | - | N/A | 0 | 7 | 1.03 |
| 132 | iRBD | Male | 66-70 | 56-60 | 8 | 18 | 2 | 18 | - | 4 | 7 | - | - | N/A | 0 | 9 | 0.80 |
| 133 | iRBD | Male | 61-65 | - | - | - | - | 18 | - | 2 | 7 | - | - | N/A | - | - | 1.74 |
| 134 | iRBD | Male | 61-65 | 61-65 | 2 | 12 | 5 | 14 | 102 | 5 | 10 | - | - | N/A | 0 | 13 | 1.19 |
| 135 | iRBD | Male | 61-65 | 46-50 | 11 | 16 | 6 | 18 | 76 | 5 | 9 | - | - | + | 0 | 12 | 1.88 |
| 136 | iRBD | Male | 61-65 | 56-60 | 4 | - | - | 11 | 9 | 2 | 5 | - | - | N/A | - | - | 0.73 |
| 137 | iRBD | Male | 66-70 | 61-65 | 3 | 12 | 3 | 13 | 2 | 5 | 7 | - | - | N/A | 0 | 4 | 2.33 |
| 138 | iRBD | Male | 61-65 | 61-65 | 1 | 15 | 2 | 14 | 9 | 0 | 3 | - | - | N/A | 0 | 5 | 2.75 |
| 139 | iRBD | Male | 61-65 | 51-55 | 10. | 25 | 1 | 15 | 6 | 3 | 5 | - | - | + | 1 | 12 | 1.99 |
| 140 | iRBD | Male | 56-60 | 51-55 | 4 | 15 | 2 | 17 | 6 | 1 | 6 | - | - | + | 0 | 3 | 1.15 |
| 141 | iRBD | Male | 61-65 | 51-55 | 8 | 13 | 9 | 11 | 8 | 3 | 0 | - | - | N/A | 0 | 11 | 1.95 |
| 142 | iRBD | Male | 56-60 | 46-50 | 12 | 22 | 2 | 14 | 5 | 2 | 9 | - | - | N/A | 0 | 11 | 1.66 |

| | | | | | | | | | | | | | | | | | |
|---|---|---|---|---|---|---|---|---|---|---|---|---|---|---|---|---|---|
| 143 | iRBD | Male | 71-75 | 61-65 | 10 | 17 | 0 | 12 | 1 | 1 | 8 | - | - | N/A | 0 | 8 | 3.43 |
| 144 | iRBD | Male | 66-70 | 51-55 | 15 | 18 | 8 | 18 | 14 | 0 | 6 | - | - | + | 0 | 10 | 3.03 |
| 145 | iRBD | Male | 56-60 | - | - | - | - | 12 | 15 | 3 | 7 | - | - | N/A | - | - | 3.95 |
| 146 | iRBD | Male | 61-65 | 51-55 | 10 | 17 | 0 | 18 | 25 | 3 | 2 | - | - | N/A | 0 | 7 | 3.27 |
| 147 | iRBD | Male | 56-60 | 46-50 | 10 | 15 | 2 | 17 | 7 | 2 | 7 | - | - | + | 0 | 2 | 2.69 |
| 148 | iRBD | Male | 71-75 | 71-75 | 3 | 19 | 1 | 18 | 18 | 9 | 9 | - | - | + | 0 | 12 | 4.28 |
| 149 | iRBD | Male | 56-60 | 51-55 | 3 | - | - | 14 | 17 | 2 | 10 | - | - | N/A | - | - | 2.51 |
| 150 | iRBD | Male | 71-75 | 56-60 | 14 | 8 | 1 | 17 | 12 | 1 | 5 | - | - | N/A | 0 | 12 | 3.41 |
| 151 | iRBD | Male | 61-65 | 61-65 | 4 | 13 | 1 | 18 | 6 | 3 | 4 | - | - | N/A | 0 | 7 | 0.84 |
| 152 | iRBD | Male | 71-75 | 66-70 | 6 | 12 | 6 | 15 | 13 | 2 | 6 | - | - | N/A | 0 | 10 | 1.39 |
| 153 | iRBD | Male | 71-75 | 56-60 | 15 | 17 | 0 | 14 | 19 | 5 | 5 | - | - | N/A | 0 | 10 | 0.52 |
| 154 | iRBD | Male | 66-70 | 61-65 | 2 | 11 | 4 | 12 | 11 | 1 | 3 | - | - | + | 0 | 6 | 1.71 |
| 155 | iRBD | Male | 76-80 | 56-60 | 20 | 19 | 0 | 14 | 15 | 7 | 6 | - | - | N/A | 0 | 7 | 0.94 |
| 156 | iRBD | Male | 66-70 | 61-65 | 5 | 18 | 6 | 15 | 40 | 2 | 4 | - | - | + | 0 | 12 | 1.17 |
| 157 | iRBD | Female | 76-80 | 66-70 | 7 | 13 | 6 | 14 | 24 | 4 | 6 | - | - | + | 1 | 7 | 1.45 |
| 158 | iRBD | Male | 71-75 | 66-70 | 5 | 17 | 1 | 14 | 16 | 2 | 0 | - | - | N/A | 0 | 11 | 1.21 |
| 159 | iRBD | Male | 66-70 | 61-65 | 5 | 13 | 4 | 15 | 21 | 7 | 4 | - | - | N/A | 0 | 10 | 0.66 |
| 160 | iRBD | Male | 56-60 | 56-60 | 1 | 11 | 3 | 16 | 29 | 1 | 4 | - | - | N/A | 1 | 13 | 0.89 |
| 161 | iRBD | Male | 61-65 | 56-60 | 2 | - | - | 18 | 15 | 2 | 8 | - | - | N/A | - | - | 1.07 |
| 162 | iRBD | Male | 56-60 | 36-40 | 19 | 17 | 0 | 15 | 38 | 3 | 7 | - | - | N/A | 0 | 12 | 0.57 |
| 163 | iRBD | Female | 56-60 | 46-50 | 6 | 13 | 4 | 17 | 27 | 2 | 9 | - | - | N/A | 1 | 12 | 1.55 |
| 164 | iRBD | Male | 66-70 | 46-50 | 20 | - | - | 11 | 40 | - | 7 | - | - | N/A | - | - | 1.66 |
| 165 | iRBD | Male | 76-80 | 66-70 | 6 | 15 | 3 | 15 | 18 | 12 | 10 | - | - | N/A | 1 | 11 | 3.78 |
| 166 | HC | Male | 21-25 | - | - | 15 | - | 15 | - | - | - | - | - | N/A | 0 | 7 | 1.04 |
| 167 | HC | Male | 16-20 | - | - | 12 | 3 | 15 | - | - | 10 | - | - | N/A | 0 | 2 | 1.84 |
| 168 | HC | Male | 66-70 | - | - | 16 | 7 | 10 | - | - | 11 | - | - | N/A | 0 | 0 | 2.15 |
| 169 | HC | Female | 56-60 | - | - | 16 | 0 | 17 | - | - | 11 | - | - | N/A | 0 | 0 | 0.72 |
| 170 | HC | Male | 66-70 | - | - | 12 | 0 | 12 | - | - | 10 | - | - | N/A | 0 | 1 | 2.68 |
| 171 | HC | Female | 51-55 | - | - | 15 | 1 | 18 | - | - | 11 | - | - | N/A | 0 | 1 | 1.34 |
| 172 | HC | Female | 16-20 | - | - | 14 | 4 | 15 | - | - | 11 | - | - | N/A | 0 | 1 | 0.82 |

| | | | | | | | | | | | | | | | | | |
|---|---|---|---|---|---|---|---|---|---|---|---|---|---|---|---|---|---|
| 173 | HC | Female | 16-20 | - | - | 14 | 5 | 16 | - | - | 10 | - | - | N/A | 0 | 3 | 0.60 |
| 174 | HC | Female | 76-80 | - | - | 12 | 4 | 18 | - | - | 11 | - | - | N/A | 0 | 3 | 1.16 |
| 175 | HC | Female | 66-70 | - | - | 14 | 0 | 17 | - | - | 11 | - | - | N/A | 0 | 3 | 2.48 |
| 176 | HC | Male | 71-75 | - | - | 18 | 0 | 14 | 27 | - | 11 | - | - | N/A | 0 | 0 | 0.72 |
| 177 | HC | Male | 71-75 | - | - | 10 | 2 | 13 | 8 | - | 10 | - | - | N/A | 0 | 0 | 1.48 |
| 178 | HC | Female | 66-70 | - | - | 11 | 0 | 18 | 9 | - | 11 | - | - | N/A | 3 | 2 | 1.11 |
| 179 | HC | Female | 61-65 | - | - | 20 | 4 | 18 | 1 | - | 11 | - | - | N/A | 0 | 2 | 0.50 |
| 180 | HC | Female | 66-70 | - | - | 18 | 7 | 18 | 10 | - | 12 | - | - | N/A | 0 | 2 | 0.38 |
| 181 | HC | Male | 31-35 | - | - | 16 | 7 | 17 | 1 | - | 11 | - | - | N/A | 0 | 1 | 0.63 |
| 182 | HC | Female | 71-75 | - | - | 18 | 1 | 18 | 10 | - | 10 | - | - | N/A | 1 | 0 | 0.71 |
| 183 | HC | Male | 76-80 | - | - | 23 | 5 | 15 | 0 | - | 11 | - | - | N/A | 0 | 2 | 0.42 |
| 184 | HC | Female | 56-60 | - | - | 16 | 0 | 18 | 18 | - | 12 | - | - | N/A | 0 | 4 | 1.33 |
| 185 | HC | Female | 46-50 | - | - | 12 | - | 17 | - | - | - | - | - | N/A | 0 | 6 | 0.52 |
| 186 | HC | Female | 61-65 | - | - | 13 | 2 | 18 | - | - | - | - | - | N/A | 1 | 2 | 0.79 |
| 187 | HC | Female | 61-65 | - | - | 14 | 2 | 17 | 3 | - | 8 | - | - | N/A | 0 | 1 | 0.71 |
| 188 | HC | Male | 36-40 | - | - | 20 | 0 | 18 | 4 | - | 11 | - | - | N/A | 0 | 1 | 1.96 |
| 189 | HC | Male | 61-65 | - | - | 13 | 1 | - | - | - | - | - | - | N/A | 2 | 2 | 0.57 |
| 190 | HC | Female | 56-60 | - | - | 20 | 0 | 18 | 18 | - | 2 | - | - | N/A | 0 | 0 | 0.55 |
| 191 | HC | Male | 51-55 | - | - | 15 | 0 | 14 | 4 | - | 10 | - | - | N/A | 0 | 0 | 1.96 |
| 192 | HC | Female | 66-70 | - | - | 19 | 2 | 15 | 1 | - | 10 | - | - | N/A | 0 | 0 | 0.68 |
| 193 | HC | Female | 61-65 | - | - | 12 | 4 | 17 | 34 | - | 11 | - | - | N/A | 0 | 0 | 0.77 |
| 194 | HC | Male | 56-60 | - | - | 16 | 1 | - | 11 | - | 11 | - | - | N/A | 0 | 2 | 0.63 |
| 195 | HC | Female | 71-75 | - | - | 16 | 11 | 18 | 15 | - | 11 | - | - | N/A | 0 | 4 | 0.48 |
| 196 | HC | Female | 66-70 | - | - | 12 | 0 | 17 | 16 | - | 6 | - | - | N/A | 0 | 4 | 0.72 |
| 197 | HC | Female | 46-50 | - | - | 17 | 1 | 18 | 1 | - | 11 | - | - | N/A | 0 | 0 | 0.58 |
| 198 | HC | Male | 61-65 | - | - | 18 | 2 | 18 | 4 | - | 11 | - | - | N/A | 0 | 2 | 0.28 |
| 199 | HC | Male | 71-75 | - | - | 22 | 1 | 13 | 14 | - | 7 | - | - | N/A | 0 | 3 | 0.49 |
| 200 | HC | Female | 46-50 | - | - | 15 | 2 | 18 | 18 | - | 11 | - | - | N/A | 0 | 5 | 0.44 |
| 201 | HC | Male | 61-65 | - | - | 16 | - | 13 | - | - | - | - | - | N/A | 0 | 9 | 0.70 |
| 202 | HC | Male | 41-45 | - | - | 21 | 0 | 14 | 0 | - | 10 | - | - | N/A | 0 | 0 | 1.02 |

| | | | | | | | | | | | | | | | | | |
|---|---|---|---|---|---|---|---|---|---|---|---|---|---|---|---|---|---|
| 203 | HC | Male | 56-60 | - | - | 22 | 0 | 15 | 3 | - | 11 | - | - | N/A | 0 | 1 | 1.25 |
| 204 | HC | Female | 66-70 | - | - | 21 | 0 | 16 | 2 | - | 7 | - | - | N/A | 0 | 0 | 0.42 |
| 205 | HC | Female | 46-50 | - | - | 13 | 3 | 17 | 3 | - | 12 | - | - | N/A | 0 | 0 | 0.48 |
| 206 | HC | Female | 36-40 | - | - | 18 | 4 | 15 | 3 | - | 11 | - | - | N/A | 0 | 0 | 0.73 |
| 207 | HC | Female | 41-45 | - | - | 17 | 3 | 18 | 21 | - | 11 | - | - | N/A | 0 | 0 | 0.85 |
| 208 | HC | Female | 26-30 | - | - | 18 | - | 18 | - | - | - | - | - | N/A | 0 | 7 | 0.94 |
| 209 | HC | Female | 51-55 | - | - | 15 | 1 | 17 | 2 | - | 11 | - | - | N/A | 0 | 3 | 0.46 |
| 210 | HC | Female | 21-25 | - | - | 18 | 6 | 17 | 18 | - | 11 | - | - | N/A | 2 | 1 | 1.60 |
| 211 | HC | Female | 51-55 | - | - | 15 | 4 | 13 | 46 | - | 11 | - | - | N/A | 0 | 4 | 1.24 |
| 212 | HC | Male | 76-80 | - | - | 12 | 0 | 13 | 32 | - | - | - | - | N/A | 0 | 5 | 2.43 |
| 213 | HC | Female | 51-55 | - | - | 13 | - | 16 | - | - | - | - | - | N/A | 3 | 7 | 0.38 |
| 214 | HC | Male | 26-30 | - | - | 13 | 0 | 18 | 2 | - | 11 | - | - | N/A | 0 | 0 | 0.45 |
| 215 | HC | Male | 36-40 | - | - | 17 | 3 | 17 | 0 | - | 12 | - | - | N/A | 0 | 3 | 0.41 |
| 216 | HC | Female | 61-65 | - | - | 23 | 5 | 18 | 16 | - | 9 | - | - | N/A | 1 | 1 | 0.72 |
| 217 | HC | Male | 61-65 | - | - | 18 | 3 | 13 | 20 | - | 11 | - | - | N/A | 0 | 2 | 0.64 |

PD = Parkinson’s disease; iRBD = isolated rapid eye movement sleep behavior disorder; HC = healthy control; CCCSS = Cleveland Clinic Constipation Scoring System; NMSS = Non-Motor Symptoms Scale; MDS-UPDRS III = Movement Disorder Society’s Unified Parkinson's Disease Rating Scale Part III; RBDSQ = iRBD screening questionnaire; N/A = not available.

**Table S2.** The coefficient of variation of each SiNaP concentration

| Concentration [fM] | Coefficient of variation [%] |
|---|---|
| 0.1 | 18.2 |
| 0.32 | 31.7 |
| 1 | 44.0 |
| 3.2 | 35.3 |
| 10 | 7.9 |
| 32 | 16.5 |
| 100 | 13.8 |
| 320 | 12.0 |
| 1000 | 10.7 |
| 3200 | 9.0 |
| Mean | 19.9 |

**Table S3.** The Spearman coefficient of correlation reveals no significant correlation between the α-synuclein aggregate concentrations in urine and other disease-relevant scores

| | PD | iRBD | HC |
|---|---|---|---|
| Age | –0.089 | 0.059 | –0,010 |
| Education | 0.028 | 0.025 | –0.253 |
| Sex | –0.166 | –0.029 | –0.135 |
| DemTect | 0.025 | –0.033 | –0.279 |
| Disease duration | 0.019 | 0.212 | N/A |
| MDS-UPDRS III | 0.079 | –0.098 | N/A |
| Screening questionnaire for parkinsonism | –0.032 | 0.094 | 0.0003 |
| RBDSQ | –0.171 | –0.050 | –0.119 |

PD = Parkinson’s disease; iRBD = isolated rapid eye movement sleep behavior disorder; HC = healthy control; MDS-UPDRS III = Movement Disorder Society’s Unified Parkinson's Disease Rating Scale Part III; N/A = not applicable; RBDSQ = REM sleep behavior disorder screening questionnaire. The Spearman coefficient of correlation was determined with GraphPad Prism Version 10.4.0.
